# Population Immunity to Hemagglutinin Head, Stalk and Neuraminidase of Highly Pathogenic Avian Influenza 2.3.4.4b A(H5N1) viruses in the United States and the Impact of Seasonal Influenza on A(H5N1) Immunity

**DOI:** 10.1101/2025.03.30.25323419

**Authors:** Zhu-Nan Li, Feng Liu, Yu-Jin Jung, Stacie Jefferson, Crystal Holiday, F. Liaini Gross, Wen-Pin Tzeng, Paul Carney, Ashley Kates, Ian A. York, Nasia Safdar, C Todd Davis, James Stevens, Terrence Tumpey, Min Z. Levine

## Abstract

The unprecedented 2.3.4.4b A(H5N1) outbreak in dairy cattle, poultry, and spillover to humans in the United States (US) poses a major public health threat. Population immunity is a critical component of influenza pandemic risk assessment. We conducted a comprehensive assessment of the population immunity to 2.3.4.4b A(H5N1) viruses and analyzed 1794 sera from 723 people (0.5-88 yrs) in multiple US geographic regions during 2021-2024. Low pre-existing neutralizing and hemagglutinin (HA) head binding antibodies and substantial cross reactive binding antibodies to N1 neuraminidase (NA) of 2.3.4.4b A(H5N1) were detected in US population. Antibodies to group 1 HA stalk were also prevalent with an age-related pattern. A(H1N1)pdm09 infection and influenza vaccination did not induce neutralizing antibodies but induced significant rise of NA inhibition (NAI) antibodies to N1 of 2.3.4.4b A(H5N1), and group 1 HA stalk antibodies. Understanding population susceptibility to novel influenza is essential for pandemic preparedness.

## Introduction

Since their emergence almost 3 decades ago^1^, highly pathogenic avian influenza (HPAI) A(H5) viruses have become endemic in wild birds and poultry in many countries, causing sporadic human infections, most from exposure to infected poultry ^2^. In recent years, spillover infections of HPAI A(H5) viruses from birds and poultry to mammals have occurred in increasing numbers of mammalian species with potential mammal-to-mammal transmission^3^. Clade 2.3.4.4b HPAI A(H5N1) viruses were first detected in wild birds in the United States (US) in 2021, and since have caused wide-spread outbreaks in US poultry ^4^. The first A(H5N1) human case in the US was identified in 2022 from direct exposure to 2.3.4.4b H5 infected poultry ^5^.

Since March 2024, an unprecedented outbreak has been ongoing among US dairy cattle in multiple states caused by 2.3.4.4b HPAI A(H5N1) viruses^6^. These viruses have transmitted among cattle, from cattle back to poultry, and spilled over into humans^6–8^. In March 2024, A(H5N1) human infection was identified in a Texas dairy farm worker after exposure to presumably infected dairy cattle, a HPAI 2.3.4.4b A(H5N1) virus was isolated, marking the first known case of cattle-to-human transmission of an avian influenza A virus^6,9^. As of March 27, 2025, 70 A(H5N1) human cases have been confirmed in multiple US states from the current outbreak. Most (65/70) were associated with exposure to infected dairy cattle or poultry, and all were caused by clade 2.3.4.4b HPAI A(H5N1) viruses^10^, raising concerns of the pandemic risk of these viruses^11^.

Influenza pandemics can occur when highly pathogenic novel influenza viruses mutate to gain sustained transmissibility among human population that has little or no pre-existing immunity. Host immunity is a critical parameter in assessing susceptibility to influenza virus infection and the risk of an influenza pandemic^11^.

Furthermore, unlike the A(H5) cases previously reported from outbreaks outside the US, which often caused severe disease with a high rate of mortality (50%)^12^, thus far, most reported A(H5N1) human cases from the current outbreak in the US have had mild clinical symptoms. Whether pre-existing host immunity plays a role in disease severity also needs to be better understood. Multiple-immune mechanisms could contribute to protection against influenza virus infection^13^; most of the past studies of human immunity to A(H5) focused on the antibodies targeting the hemagglutinin (HA) head of the viruses. Little is known about the population immunity to other immunological targets of the A(H5) viruses that could also contribute to protection and attenuation of disease, including antibodies to neuraminidase (NA) and the HA stalk.

In this study, we conducted a comprehensive assessment of the levels of neutralizing antibodies, HA head binding antibodies, functional NA inhibition antibodies (NAI), NA binding antibodies to HPAI clade 2.3.4.4b A(H5N1) viruses, and antibodies to group 1 HA stalk using 1794 sera collected from 723 participants (aged 0.5- 88 yrs) from multiple geographic regions in the US across recent influenza seasons (2021-2024). We analyzed whether seasonal influenza A virus infection or seasonal influenza vaccination can induce cross-reactive antibodies to 2.3.4.4b A(H5N1) viruses to investigate the plausible sources of the pre-existing immunity to A(H5N1) viruses.

## Results

### Low prevalence of antibodies to the HA head of clade 2.3.4.4b A(H5N1) viruses in the US

We have been monitoring the population immunity in the US to both seasonal and novel influenza viruses in a multi-season longitudinal study. From November 2021 to February 2023, 489 participants from all 10 US Department of Health and Human Services (HHS) regions (Fig 1a and Table 1) and 8 age groups (0-10 yrs, 11-20 yrs, 21-30 yrs, 31-40 yrs, 41-50yrs, 51-60 yrs, 61-70 yrs and > 70 yrs, Fig 1b) were enrolled and provided sera at 3 waves (time points) across the 2021-23 influenza seasons (Table 1 and Fig 1c-e). A total of 1467 sera were analyzed by a 12-plex multiplex influenza antibody detection assay (MIADA) (Extended Table S1). High levels of binding antibodies to the HA head of a representative seasonal A(H1N1)pdm09 virus A/Wisconsin/588/2019 were detected across most age ranges (Fig 1 c-e). In contrast, there were very low levels of pre-existing binding antibodies to the HA head of the HPAI A(H5N1) 2.3.4.4b virus A/American wigeon/South Carolina/22-000345- 001/2021 (AW/2021) during 2021-23 influenza seasons (Fig 1 c-e).

**Fig. 1.**
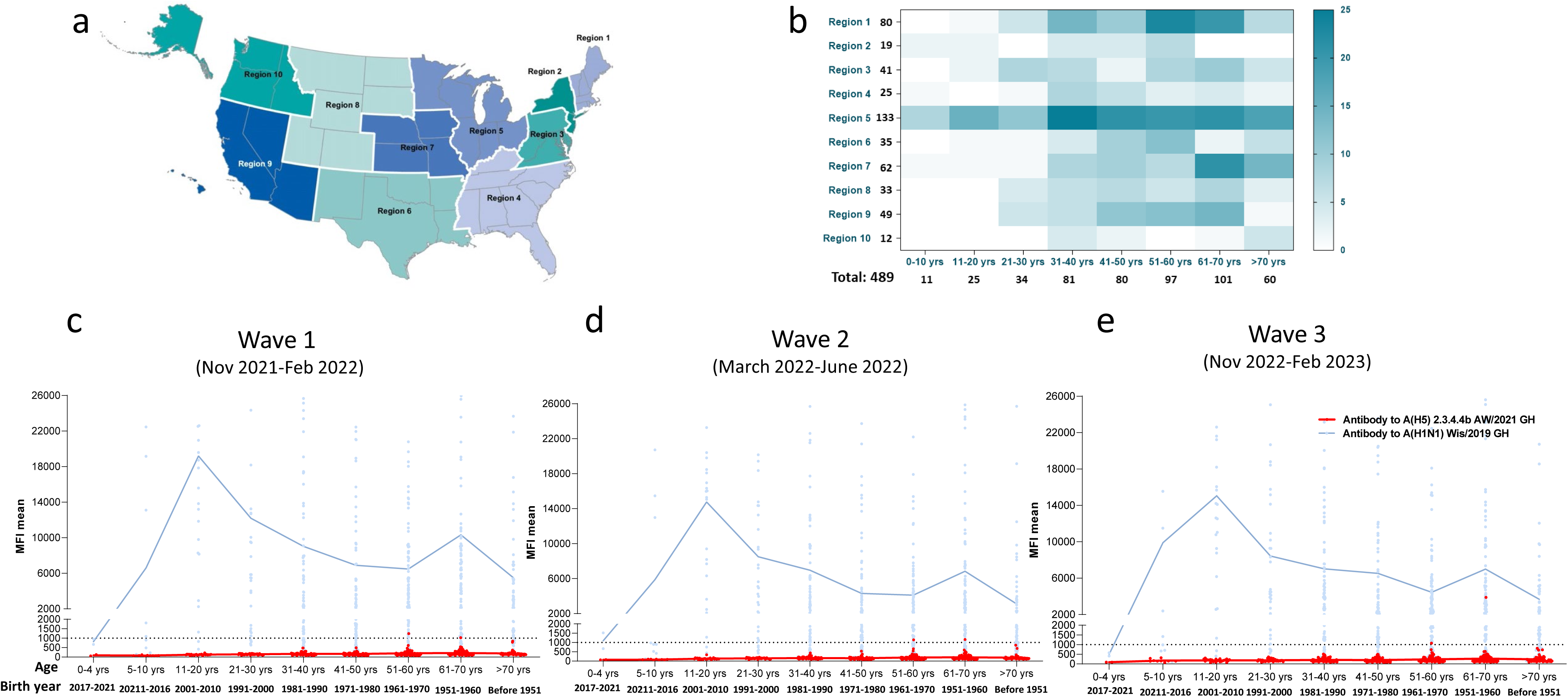
Age-stratified prevalence of binding antibodies to the hemagglutinin (HA) head of 2.3.4.4b HPAI A(H5N1) and A(H1N1)pdm09 viruses in the United States in 2021-23. (a). **Ten HHS regions in the US. (b)** Distribution of the regions and ages of 489 participants enrolled in the longitudinal population immunity study, 3 waves (time points) of sera were collected from each participant from 2021 to 2023 influenza seasons. **(c-e)** Scatter plots of age-stratified prevalence of binding antibodies to the HA head of 2.3.4.4b A(H5N1) A/American wigeon/South Carolina/22-000345-001/2021 (AW/2021) and A/Wisconsin/588/2019 H1N1pdm09 (Wis/2019) by waves (total 1467 sera collected from 489 individuals from 3 waves). Connected lines represent mean antibody MFI of each age group, dotted lines represent MFI of 1000. **(c)** Wave 1 (Nov 2021-Feb 2022) n=489; **(d)** Wave 2 (March 2022-June 2022) n=489; **(e)** Wave 3 (Nov 2022-Feb 2023) n=489.

**Table 1.**
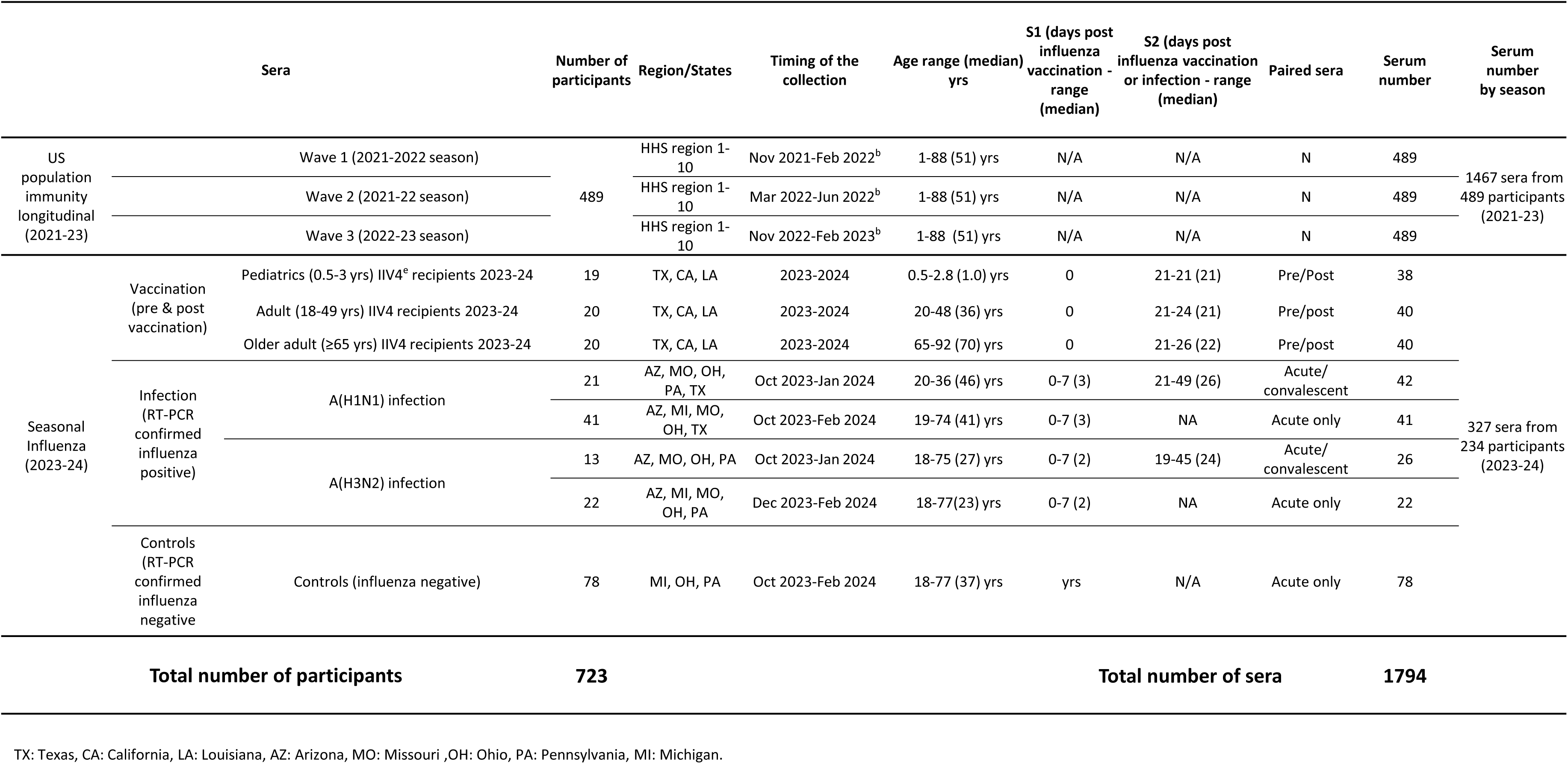
Characteristics of the participants and sera evaluated in the study.

Based on the World Health Organization (WHO) A(H5) case definition^14^, seropositivity to A(H5) virus using a single serum sample was defined as a neutralizing antibody titer ≥40 to an A(H5) virus that is antigenically similar to the viruses that are causing the current outbreak, and either hemagglutination inhibition (HI) titer to an antigenically similar A(H5) virus ≥40, or a positive result from an A(H5)-specific immunological assay such as a multiplex antibody binding assay such as MIADA. We analyzed the sensitivity and specificity of the MIADA assay in detecting HA head binding antibodies to 2.3.4.4b AW/2021 A(H5N1) using A(H5) positive sera with MN titer ≥40 to the wild-type 2.3.4.4b A(H5N1) AW/2021 virus (data not shown) and 1467 sera from the 2021-23 population immunity study collected before the current A(H5N1) outbreak as negative sera (Figure 1 c-e). At median fluorescence intensity (MFI) cutoff of 1000 to 2.3.4.4b A(H5N1) AW/2021GH, the assay had the highest J-index of 0.996 (sensitivity[100%] + specificity[99.6%]-1). To confirm seropositivity, samples from the 2021-23 population immunity study (Fig 1) that had the highest MFI (>650) values to 2.3.4.4b A(H5N1) AW/2021 GH were tested by both MN and HI assays against wild type 2.3.4.4b HPAI A(H5N1) AW/2021 virus and 2.3.4.4b HPAI A(H5N1) A/Texas/37/2024 virus. All sera were negative against both wild type A(H5N1) viruses in both assays (Extended Table S2) with no seropositivity among the population immunity sera surveyed across 3 time points during 2021-23 seasons (Fig 1).

Since the onset of the A(H5N1) outbreak in dairy cattle in early 2024, we updated the MIADA assay to a high throughput 28-plex with an expanded antigen cocktail including HA and NA of the virus isolated from 2.3.4.4b HPAI A/Texas/37/2024 (H5N1) for antibody landscape analysis (antigens shown in Extended Table S1), and assessed recent sera collected in the 2023-24 influenza season (327 sera collected from 234 participants aged 0.5-77 yrs) (Table 1). Similar to data from previous seasons, all 2023-24 season sera were low for total binding antibody to 2.3.4.4b A(H5N1) AW/2021 GH (MFI <1000, Fig 2b), MFIs to A/Texas/37/2024 (H5N1) GH were also low (<1000, Fig 2a) across all age groups, suggesting the population immunity to the HA head of the 2.3.4.4b viruses remained low in the 2023-24 season. As expected, binding antibodies to the representative seasonal A(H1N1)pdm09 virus A/Wisconsin/67/2022 GH were high across most age groups (Fig 2a-b).

**Fig. 2.**
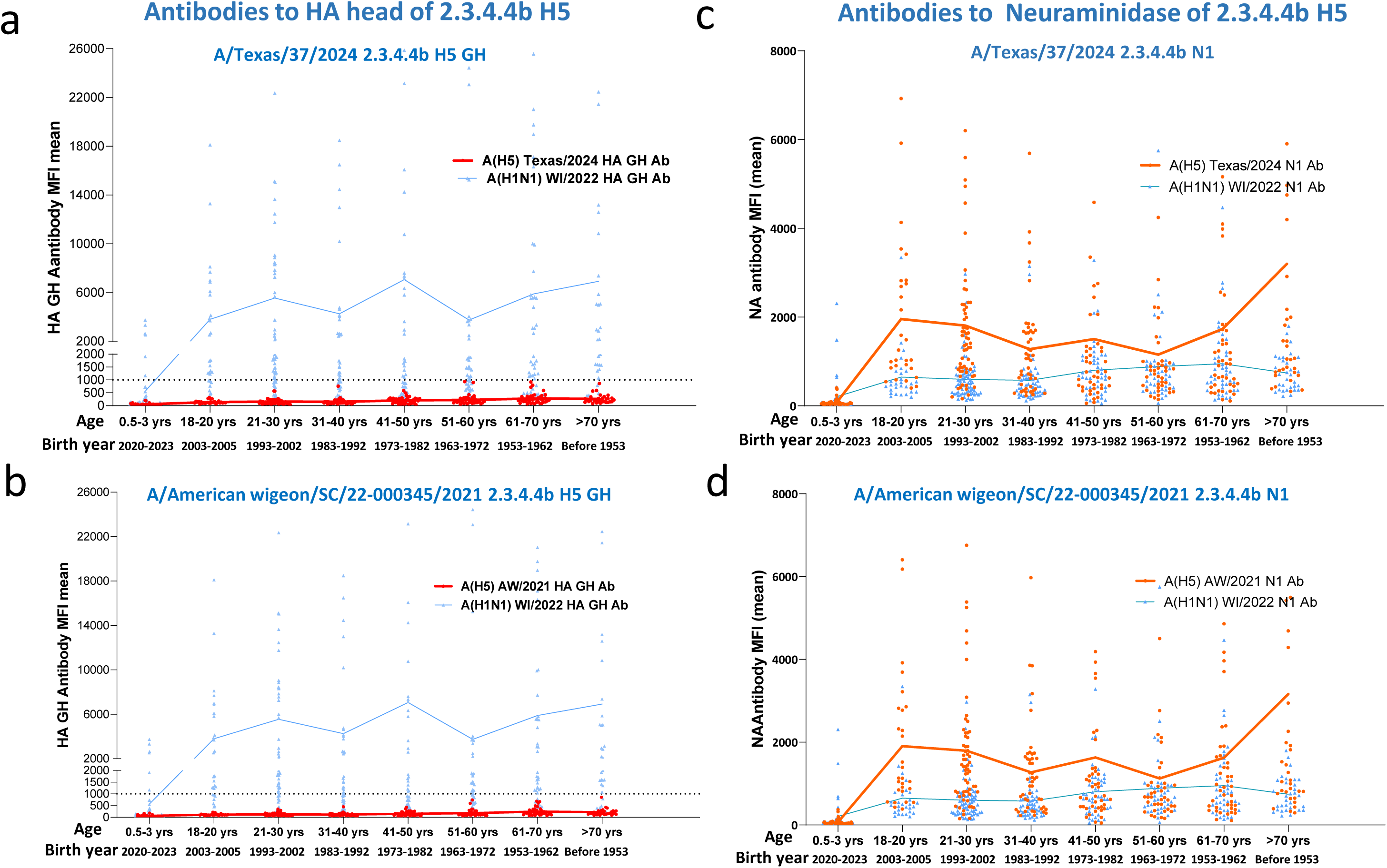
Age-stratified prevalence of binding antibodies to the hemagglutinin (HA) head, neuraminidase of HPAI 2.3.4.4b A(H5N1) and A(H1N1)pdm09 viruses in the US population in 2023-24. Total 327 sera collected from 234 participants during 2023-24 influenza season were stratified into 8 age groups. Age-stratified scatter plot of binding antibodies: **(a)** Antibodies to HA globular head (GH) of 2.3.4.4b A(H5N1) A/Texas/37/2024 and A(H1N1) pdm09 A/Wisconsin/67/2022 (WI/2022) **(b)** Antibodies to HA GH of 2.3.4.4b A/American wigeon/SC/22-000345/2021 (AW/2021) and A(H1N1)pdm09 WI/2022; **(c)** Antibodies to N1 neuraminidase of 2.3.4.4b A(H5N1) A/Texas/37/2024 and A(H1N1) pdm09 WI/2022; **(d).** Antibodies to N1 neuraminidase of 2.3.4.4b AW/2021 and A(H1N1)pdm09 WI/2022. Connected lines represent mean antibody MFI of each age group, dotted line in (a) and (b) represent MFI of 1000.

### Seasonal influenza virus infection and vaccination did not induce neutralizing antibodies to 2.3.4.4b virus or binding antibodies to 2.3.4.4b A(H5N1) virus HA head

Next, we assessed whether seasonal influenza A virus infection or influenza vaccination could induce cross-reactive antibodies to 2.3.4.4b A(H5N1) viruses. We collected acute (within 7 days from symptom onset) and convalescent (19-49 days since symptom onset) sera from reverse transcription polymerase chain reaction (RT-PCR) confirmed influenza A(H1N1)pdm09 and A(H3N2) virus infections in the 2023-24 season and analyzed both neutralizing and binding antibody responses to seasonal and A(H5N1) viruses. Seasonal influenza A virus infection induced a significant rise of neutralizing antibodies to the infecting A(H1N1)pdm09 (Fig 3a) or A(H3N2) (Fig 3b) viruses, but no neutralizing antibodies (MN<10) to AW/2021 2.3.4.4b A(H5N1) virus were detected in any of the acute and convalescent sera (Fig 3 a-b). We also analyzed pre- and post- seasonal influenza vaccine sera collected from older adults (≥ 65 yrs), adults (18-49 yrs), and pediatric (0.5-3 yrs) populations who received 2023-24 inactivated quadrivalent influenza vaccines (IIV4). IIV4 seasonal influenza vaccination induced a significant rise in neutralizing antibodies to A(H1N1)pdm09 vaccine virus, but no neutralizing antibodies (MN<10) to 2.3.4.4b AW/2021 H5N1 virus were detected in all pre- and post-vaccination sera from all age groups (Fig 3 c-e).

**Fig. 3.**
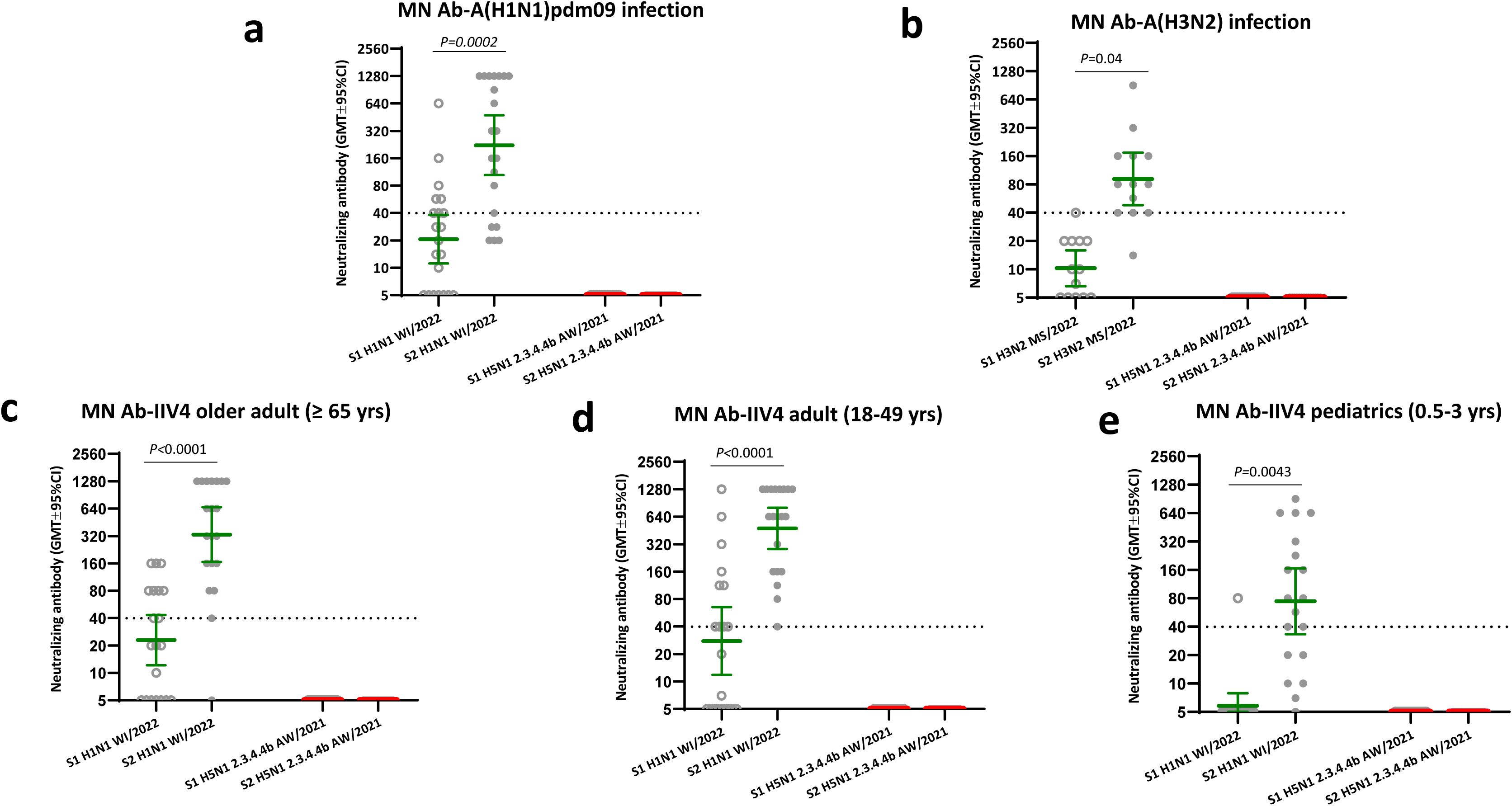
Seasonal influenza infection and vaccination in 2023-24 did not induce neutralizing antibodies to 2.3.4.4b HPAI A(H5N1) virus. Paired serum samples collected from seasonal influenza infection and quadrivalent inactivated influenza vaccination (IIV4) participants during 2023-24 influenza season were tested by microneutralization (MN) assays. Neutralizing antibody titers against 2.3.4.4b A(H5N1) A/American wigeon/South Carolina/22-000345/2021 (AW/2021) candidate vaccine virus (CVV) and seasonal influenza A(H1N1) pdm09 A/Wisconsin/67/2022 (WI/2022) were plotted in scatter plots. **(a)** Acute (S1) and convalescent (S2) sera from A(H1N1)pdm09 infection (n=21); **(b)** Acute (S1) and convalescent (S2) sera from A(H3N2) infection (n=13); **(c)** Pre (S1) and post (S2) vaccination sera collected from older adults (≥ 65 yrs) who received IIV4 (n=20); **(d)** Pre (S1) and post (S2) vaccination sera collected from adults (18-49 yrs) who received IIV4 (n=20); (**e)** Pre (S1) and post (S2) vaccination sera collected from pediatric participants (0.5-3 yrs) who received IIV4 (n=19). Y axis: geometric mean titers (GMT) and 95% confidence interval of neutralizing antibodies. Dashed line: Neutralizing antibody titer of 40 threshold. Statistical analyses were conducted using two tailed paired t test, *p*<0.05 is considered statistically significant. \**p*<0.05.

In the HA binding antibody landscape analysis by the MIADA assay, seasonal A(H1N1)pdm09 or A(H3N2) virus infection or IIV4 vaccination in 2023-24 not only induced significant (*p*<0.05) rise of binding antibodies to the HA head of the seasonal virus of exposure (either the infecting virus or antigens in the quadrivalent vaccines), in most cases it also back-boosted antibodies to older seasonal viruses within the same subtype (Fig 4). None, however, induced a significant rise in binding antibodies to the HA head of either AW/2021 or A/Texas/37/2024 2.3.4.4b HPAI A(H5N1) viruses (Fig 4a-e).

**Fig. 4.**
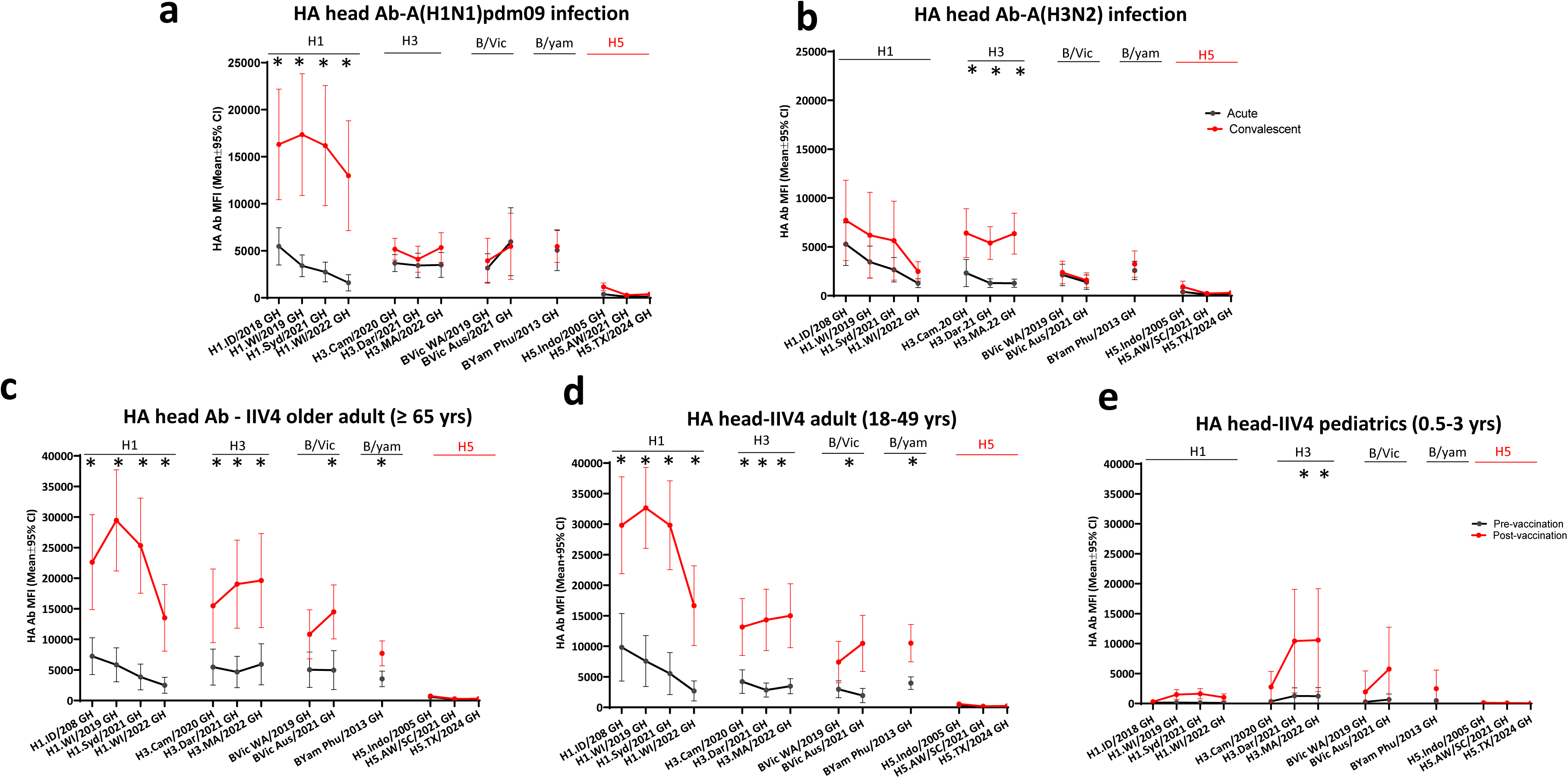
Seasonal Influenza infection and vaccination in 2023-24 did not significantly induce binding antibodies to the HA Head of A(H5) 2.3.4.4b viruses. Paired serum samples collected from seasonal influenza infection and quadrivalent inactivated influenza vaccination (IIV4) participants during 2023-24 influenza season were tested by 28-Plex MIADA, binding antibody levels (MFI) to HA head were plotted. **(a)** Acute (S1) and convalescent (S2) sera from A(H1N1)pdm09 infection (n=21); **(b)** Acute (S1) and convalescent (S2) sera from A(H3N2) infection (n=13); **(c)** Pre (S1) and post (S2) vaccination sera collected from older adults (≥ 65 yrs) who received IIV4 (n=20); **(d)** Pre (S1) and post (S2) vaccination sera collected from adults (18-49 yrs) who received IIV4 (n=20); (**e)** Pre (S1) and post (S2) vaccination sera collected from pediatric participants (0.5-3 yrs) who received IIV4 (n=19). Y axis: Mean MFIs and 95% confidence interval (CI). Statistical analyses were conducted using two tailed paired t test, *p*<0.05

### Pre-existing functional NAI antibodies and binding antibodies to the N1 NA of 2.3.4.4b A(H5N1) virus in the US population from past exposure to seasonal A(H1N1)pdm09 viruses

We analyzed the prevalence of antibodies to the N1 NAs of two 2.3.4.4b viruses in sera collected from 2023-24 participants (327 sera from 234 participants). Surprisingly, there were substantial levels of pre-existing total binding antibodies to the N1 NAs of both 2.3.4.4b A(H5N1) viruses (AW/2021 and A/Texas/37/2024) across most age groups >18 yrs. The youngest children (0.5-3 yrs) had the lowest and older adults (> 70 yrs) had the highest pre-existing binding antibodies to the N1 NA of 2.3.4.4.b A(H5N1) (Fig 2 c-d).

We then investigated whether the high levels of pre-existing antibodies to N1 of A(H5N1) were due to cross-reactivity from seasonal influenza virus A virus infection or vaccination. NA binding antibody landscape analysis using the acute and convalescent sera collected from seasonal influenza infections showed that A(H1N1)pdm09 virus infection in the 2023-24 season not only boosted N1 binding antibodies to recent circulating seasonal A(H1N1)pdm09 viruses, it also induced significant rises in N1 binding antibodies to both 2.3.4.4b A(H5N1) viruses (AW/2021 and A/Texas/37/2024) (*p*<0.05) (Fig. 5a), whereas A(H3N2) virus infection did not boost significant rises in N1 binding antibodies to A(H5N1) (p>0.05) (Fig 5b). Following seasonal IIV4 vaccination in 2023-24 in 3 age groups (older adult ≥65 yrs, adult 18-49 yrs, and pediatric 0.5-3 yrs) (Fig 5 c-e), there were some rises of N1 binding antibodies to 2.3.4.4b A(H5N1) viruses in adult and older adult groups, but these were not statistically significant (Fig 5 c-d).

**Fig. 5.**
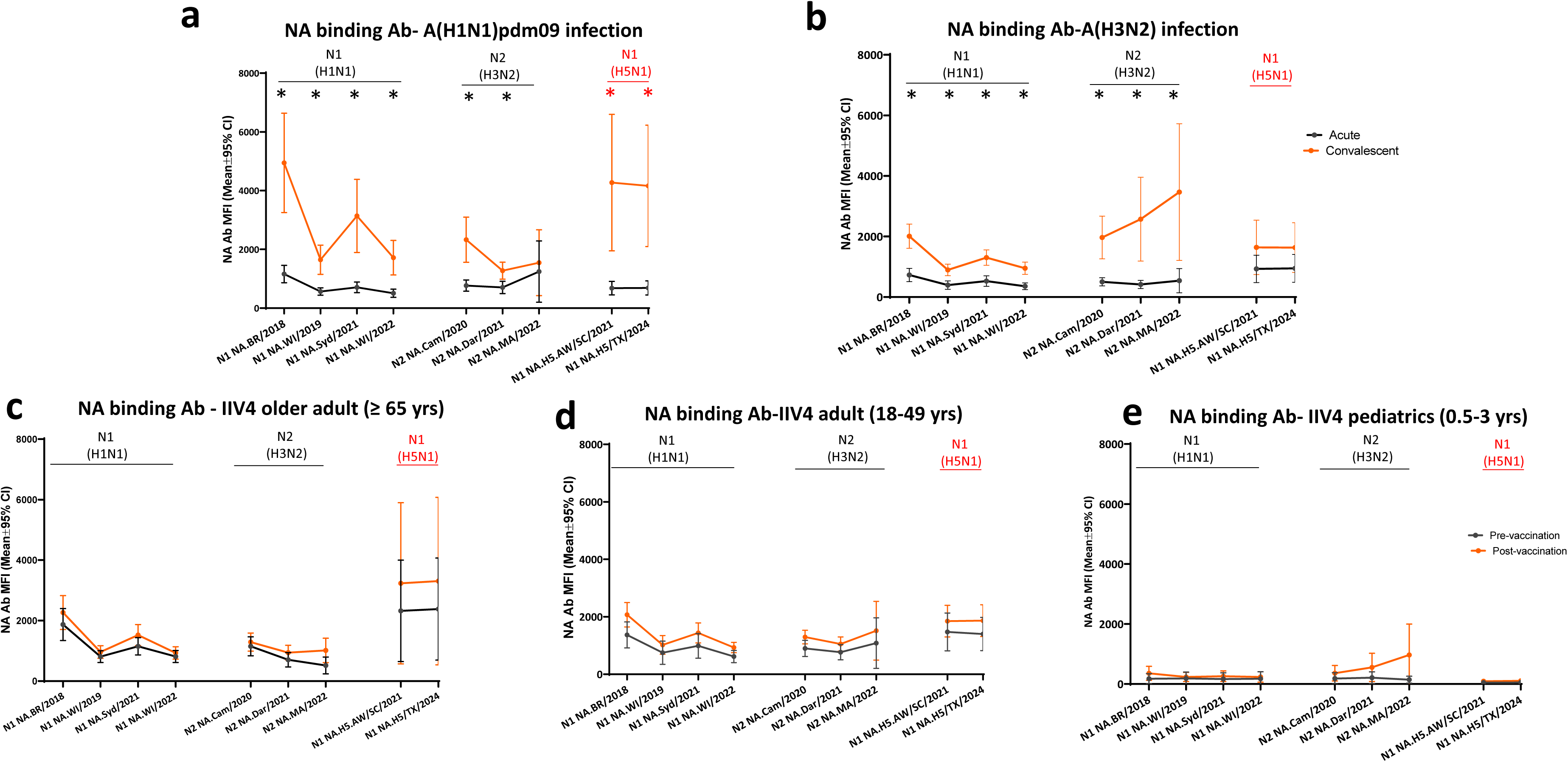
Seasonal A(H1N1)pdm09 influenza virus infection, but not A(H3N2) virus infection or seasonal vaccination induced significant rise of binding antibodies to the N1 neuraminidase of A(H5N1) **2.3.4.4b.** Paired serum samples collected from seasonal influenza infection and quadrivalent inactivated influenza vaccination (IIV4) vaccination participants during 2023-24 influenza season were tested by 28-Plex MIADA, binding antibody levels (MFI) to NA were plotted. **(a)** Acute (S1) and convalescent (S2) sera from A(H1N1) infection (n=21); **(b)** Acute (S1) and convalescent (S2) sera from A(H3N2) infection (n=13); **(c)** Pre (S1) and post (S2) vaccination sera collected from older adults (≥ 65 yrs) who received IIV4 (n=20); **(d)** Pre (S1) and post (S2) vaccination sera collected from adults (18-49 yrs) who received IIV4 (n=20); (**e)** Pre (S1) and post (S2) vaccination sera collected from pediatric participants (0.5-3 yrs) who received IIV4 (n=19). Y axis: mean MFIs and 95% confidence interval (CI). Statistical

Next, to assess the functional NAI antibodies, we constructed H6Nx reverse genetic viruses with mismatched H6 (to eliminate potential interference from HA antibodies in the human sera) and NA from seasonal A(H1N1)pdm09 (A/Wisconsin/588/2019), A(H3N2) (A/Darwin/6/2021), and A(H5N1) (N1 from HPAI 2.3.4.4b A/Texas/37/2024) viruses to assess the NAI antibody responses from seasonal influenza exposure by Enzyme-linked lectin assay (ELLA). 2023-24 seasonal A(H1N1)pdm09 virus (with N1 NA) infection, but not A(H3N2) (with N2 NA) infection, induced a significant rise of functional cross-reactive NAI antibodies targeting N1 of 2.3.4.4b A/Texas/37/2024 (H5N1) virus (Fig 6 a-b). Furthermore, 2023-24 seasonal influenza IIV4 vaccination also induced significant NAI antibody responses to N1 of 2.3.4.4b A/Texas/37/2024 A(H5N1) virus in older adult (≥ 65 yrs), adult (18-49 yrs), and pediatric (0.5-3 yrs) participants (Fig 6 c-e).

**Fig. 6:**
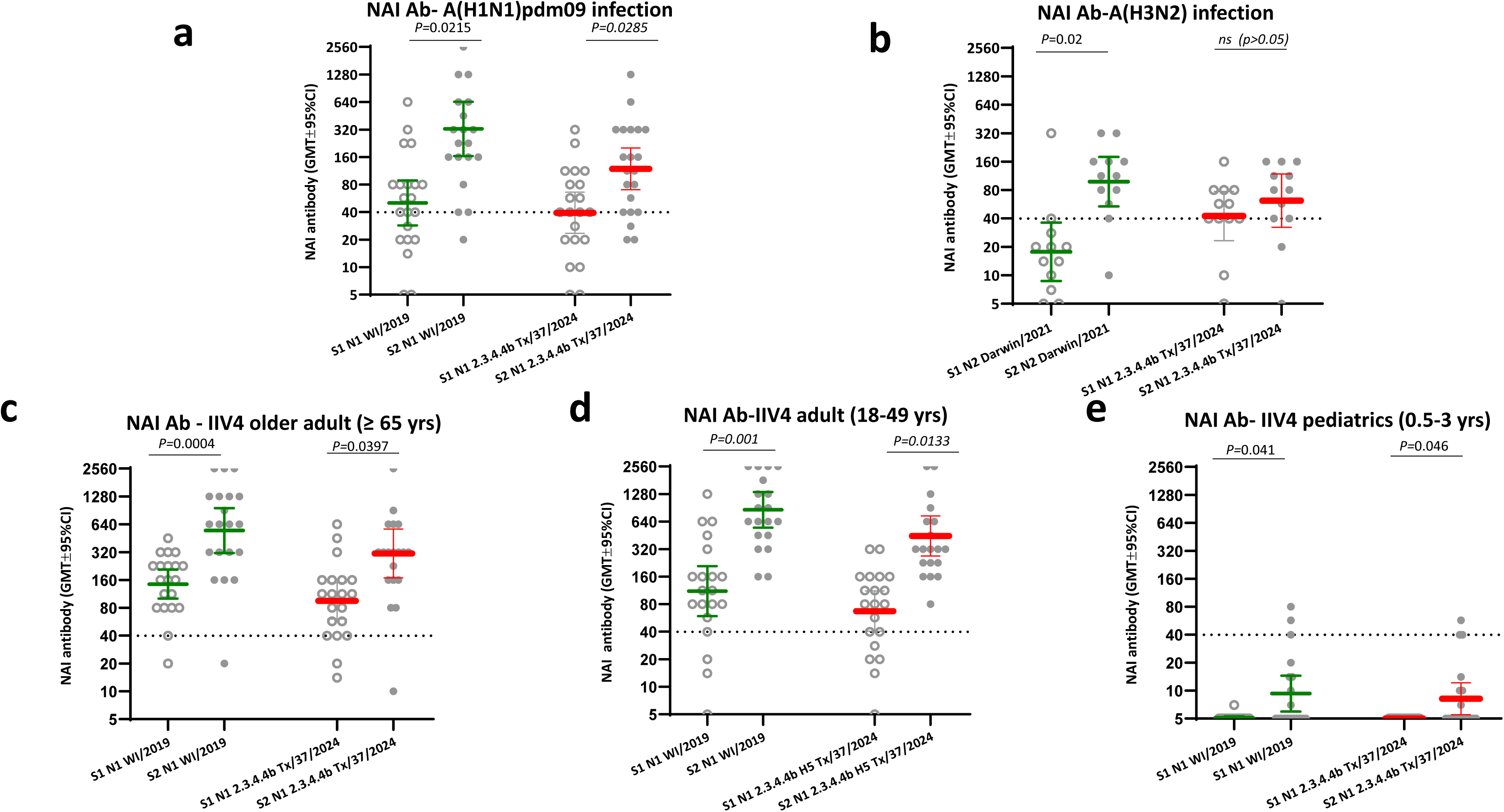
A(H1N1)pdm09 virus infection and seasonal influenza vaccination in 2023-24 induced significant rise of functional neuraminidase inhibition antibody (NAI) responses to N1 of 2.3.4.4b HPAI A(H5N1) virus. Paired serum samples collected from seasonal influenza A infection and quadrivalent inactivated influenza vaccination (IIV4) vaccination participants during 2023-24 influenza season were tested by ELLA assay using H6Nx reverse genetic viruses with mismatched HA and target NAs. Functional neuraminidase inhibition antibody (NAI) to N1 of 2.3.4.4b A(H5N1) A/Texas/37/2024, N1 from seasonal A(H1N1) and N2 neuraminidase from seasonal A(H3N2) were plotted in scatter plots. **(a)** NAI antibodies in acute (S1) and convalescent (S2) sera from A(H1N1)pdm09 infection (n=21); (b) NAI antibodies in acute (S1) and convalescent (S2) sera from A(H3N2) infection (n=12); **(c)** NAI antibodies in pre (S1) and post (S2) vaccination sera collected from older adults (≥ 65 yrs) who received IIV4 (n=20); **(d)** NAI antibodies in pre (S1) and post (S2) vaccination sera collected from adults (18-49 yrs) who received IIV4 (n=20); (**e)** NAI antibodies in pre (S1) and post (S2) vaccination sera collected from pediatric participants (0.5-3 yrs) who received IIV4 (n=19). Y axis: geometric mean titers (GMT) of NAI antibodies with 95% confidence interval (CI). Statistical comparisons were conducted using two tailed paired t test, *p*<0.05 is considered statistically significant. \**p*<0.05.

### Prevalence of antibodies to the group 1 HA stalk in the US population and the impact of seasonal influenza vaccination and infection on antibodies to the group 1 HA stalk

Influenza viruses are classified into two groups based on their antigenic properties of HA: group 1 and group 2. Both A(H5) and A(H1) belong to group 1 HA. HA stalks from A(H5N1) and A(H1N1)pdm09 are highly conserved (Extended Fig 1) and cross-reactive^15^.

We assessed the prevalence of antibodies to the group 1 HA stalk (headless H1 stalk from A/Michigan/15/2015 H1N1). There was a wide range of pre-existing anti group 1 HA stalk antibodies in the population (Fig 7a). In sera collected in 2023-24, when stratified by age groups with 2–10-year age intervals, the mean MFI to group 1 HA stalk antibody in each age group increased with age, with the lowest prevalence of group1 HA stalk antibody in very young children (0.5-3 yrs) and highest in older adults >70 yrs, reflecting the accumulation of past exposures to group 1 influenza viruses (Fig 7a).

**Fig. 7.**
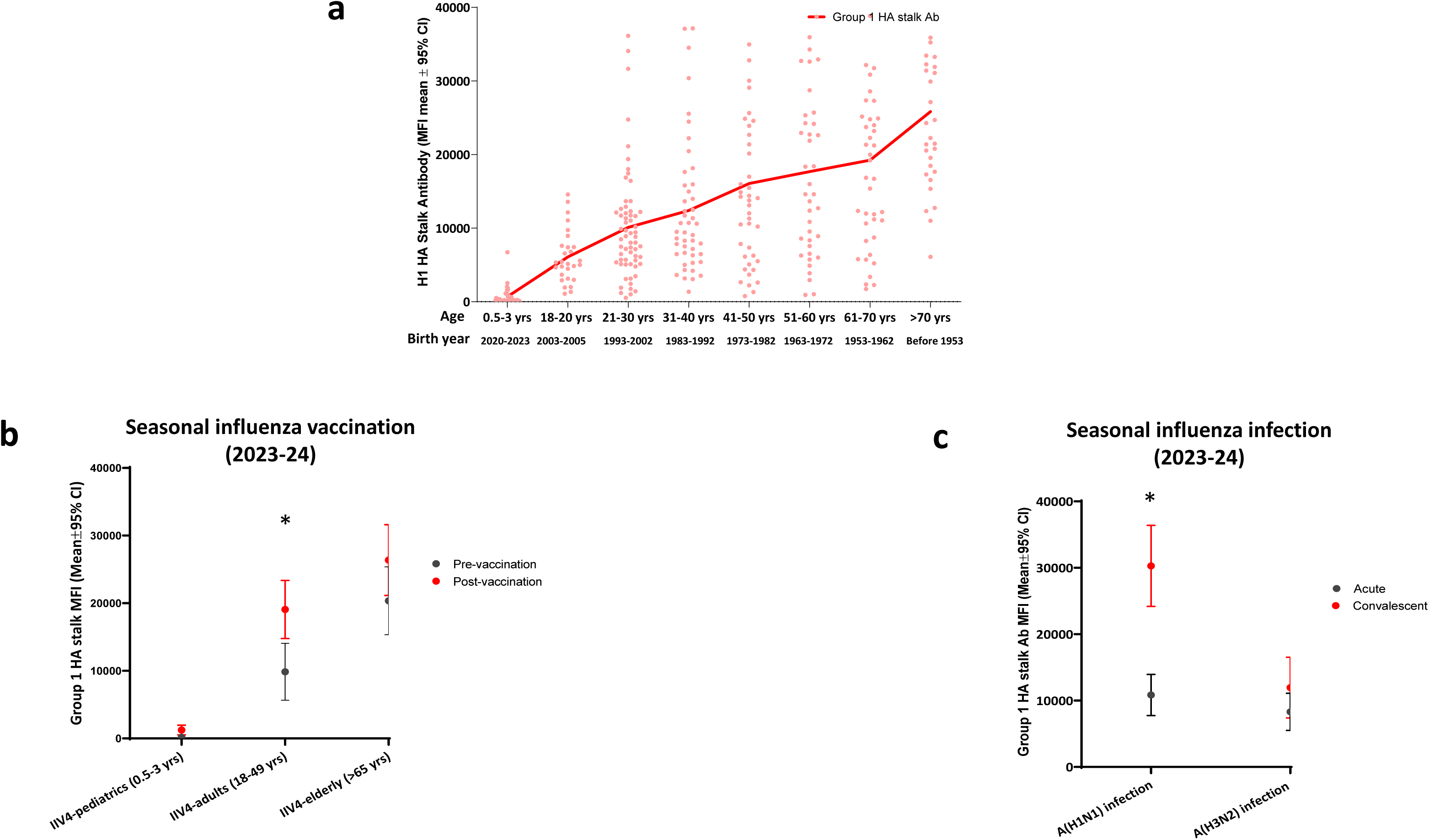
Prevalence of antibodies to the group 1 HA stalk in the US population, and antibody responses to group 1 HA talk from seasonal influenza virus infection and vaccination. **(a)** Age-stratified scatter plot of antibodies to group 1 HA stalk in 2023-24, 327 sera were collected from 234 participants from 8 age groups. Connected lines represent mean antibody MFI of each age group. **(b)** Group 1 HA stalk antibodies in pre (S1) and post (S2) vaccination sera collected in 2023-24 from quadrivalent inactivated influenza vaccination (IIV4) in older adult (≥ 65 yrs) (n=20), adult (18-49 yrs) (n=20), and pediatric participants (0.5-3 yrs) (n=19). **(c)** Group 1 HA stalk antibodies in acute (S1) and convalescent (S2) sera from A(H1N1)pdm09 infection (n=21) and A(H3N2) infection (n=13) in 2023-24 season. Y axis: mean MFIs and 95% confidence interval (CI). Statistical analyses were conducted using two tailed paired t test, *p*<0.05 is considered statistically significant. \**p*<0.05.

Lastly, to elucidate the source of the anti HA stalk antibodies, we analyzed antibody responses to group 1 HA stalk following seasonal influenza vaccination and infection. In the 3 age groups that received 2023-24 IIV4 vaccination, pre-existing group 1 HA stalk antibody levels increased with age, young children (0.5-3 yrs) had the lowest and older adults (>65 yrs) had the highest antibodies to the group 1 HA stalk (Fig 7b). IIV4 vaccination elicited significant rise of group 1 HA stalk antibodies in adults (18-49 yrs) (*p*<0.05) (Fig 7b). Furthermore, adults infected with A(H1N1)pdm09 (group 1 virus) (*p*<0.05), but not those infected with A(H3N2) (group 2 virus) (*p*>0.05), mounted significantly rise of group 1 HA stalk antibody responses (Fig. 7c). Of note, A(H1N1)pdm09 infection induced higher rise of group 1 HA stalk antibodies (mean fold rise in convalescent/acute sera: 2.8) than IIV4 vaccination (mean fold rise in post/ pre-vaccination sera: 1.9) (Fig 7b-c) in adults.

## Discussion

Population immunity against new emerging novel viruses is a key factor for influenza pandemic risk assessment^16^. Amid the ongoing A(H5N1) outbreaks in cattle and poultry and the continued spillover to humans, our study provides a timely and comprehensive assessment of the population immunity in the US to 2.3.4.4b A(H5N1) viruses. Results from the current study demonstrate that the levels of the pre-existing neutralizing antibodies and the HA head binding antibodies to 2.3.4.4b A(H5) viruses in the US population are low, consistent with previous reports of low seroprevalence (mostly measured by HI antibodies) even in populations at increased risk of A(H5) exposure (e.g., poultry workers)^12^. However, our study revealed that the population in the US was not completely immunologically naive to the 2.3.4.4b A(H5N1) viruses: there were substantial levels of pre-existing antibodies to the N1 NA of 2.3.4.4b A(H5N1) virus, and group 1 HA stalk antibodies in an age-related pattern. Furthermore, these pre-existing cross-reactive immunities to A(H5N1) virus (group 1) were mostly likely from past exposures to seasonal A(H1N1)pdm09 (group 1), not A(H3N2) (group 2) viruses.

While neutralizing antibodies targeting the HAs of the influenza virus are the main correlate of protection in reducing the risk of influenza virus infections, multiple immune mechanisms can contribute to protection from influenza^13^. Although seasonal influenza A(H1N1)pdm09 virus infection and influenza vaccination did not induce neutralizing and HA head binding antibodies to A(H5N1) viruses (Fig 3-4), both could induce significant rise of cross-reactive functional NAI antibodies to the N1 NA of 2.3.4.4b A(H5N1) (Fig 6). Sequence analysis showed that there is significant genetic distance between the HA head of the 2.3.4.4b A(H5N1) and A(H1N1)pdm09 viruses, with amino acid homology at approximately only 53%, and differences across multiple antigenic sites (Extended Table S3 and Extended Fig 2). In contrast, there is a higher level of amino acid sequence homology (86-88%) between the N1 NA sequences of 2.3.4.4b A(H5N1) and recent circulating seasonal A(H1N1)pdm09 viruses (Extended Table S4).

NA antibodies have been considered as an independent correlate to protect against influenza^17^. Although they cannot prevent infection, NA antibodies can prevent virus egress, reduce viral shedding, thus could attenuate disease and lessen disease severity ^18,19^. Past studies have demonstrated that seasonal influenza virus infection and vaccination could induce cross-reactive and protective NA-specific antibodies, conferring prophylactic protection against older avian A(H5N1) viruses in mice, ferret models and humans^20^ ^21^ ^22^. There were 10 amino acid differences between the N1 NA of 2.3.4.4b A/Texas/37/2024 H5N1 and A/Wisconsin/67/2022 (H1N1)pdm09 viruses among the putative NA antigenic sites (Extended data Fig 3 and 4). A previous epitope mapping study showed that three primary amino acids on N1 NA (G249, N273 and N309) were targeted by the NA-reactive antibodies ^23^, and all these three amino acids are conserved among the N1 NAs between 2.3.4.4b A(H5N1) and recent A(H1N1)pdm09 viruses (Extended Fig 3). Additional NA epitopes, such as S95 and D451 which were shown to play a role in the inhibition of A(H1N1)pdm09 virus ^24^, were also conserved among the N1 NAs investigated in this study (Extended Fig 3). Past exposure to seasonal A(H1N1)pdm09 viruses could induce cross-reactive functional NAI antibodies to the N1 NA 2.3.4.4b A(H5N1) virus. To date, the threshold of the NAI antibody that correlates with protection still remains to be determined, the amount of NA in the US licensed seasonal vaccines is not regulated, thus the level of cross-reactive NAI antibodies induced by seasonal vaccination may be highly variable. Nevertheless, the role of NA antibodies in influenza protective immunity has been well recognized. Whether these pre-existing NAI antibodies could have contributed to the mild clinical outcomes in most of the human A(H5) virus infections detected thus far during the current outbreak warrants further investigation.

Broadly cross-reactive HA stalk antibodies have been considered as one of the targets for universal influenza vaccine because HA stalk is highly conserved^25,26^. Our results suggested that the group 1 HA stalk antibodies exist in the US population in an age-related pattern: they increased with age and birth cohorts, and can be further boosted upon A(H1N1)pdm09 infection and IIV4 vaccination in adults (Fig 7). Both A(H1) HA and A(H5) HA belong to the same phylogenetic group 1, and the HA stalks are fairly conserved between the A/Texas/37/2024 H5N1 and A/Michigan/45/2015 H1N1 viruses (Extended Fig 1). Gostic et al. suggested through statistical modeling that childhood imprinting with A(H1N1) virus could provide protection for A(H5N1) viruses (belong to the same phylogenetic group 1) through conserved HA epitopes ^27^. Stalk antibodies are thought to offer protection through Fc receptor-dependent mechanisms such as antibody dependent cellular cytotoxicity (ADCC). Whether and to what extent these pre-existing group 1 HA stalk antibodies can confer protection to A(H5N1) virus infection in humans still needs to be better understood.

Vaccination is the most effective public health measure to prevent influenza and annual seasonal influenza vaccination is recommend for people aged ≥6 months in the United States^28^. Influenza vaccines are also anticipated to be used as a medical countermeasure in the event of an influenza pandemic^29^. Our recent study in the ferret model suggested that prior influenza A(H1N1)pdm09 virus infection may confer some level of protection against influenza A(H5N1) clade 2.3.4.4.b virus^30^. While seasonal influenza vaccines are not intended to prevent A(H5) influenza, our data from humans revealed that they could induce cross-reactive and potentially cross-protective NA and HA stalk antibodies; the clinical implications of these responses in A(H5) human infection merit further study.

During a novel influenza A virus outbreak, timely serologic studies can detect both symptomatic and asymptomatic infections and provide more robust estimates of total infections within a population ^31^. When using a single convalescent serum, the seropositivity criteria in the WHO A(H5) case definition ^14^ requires microneutralization titers ≥40, and either HI ≥40, or a positive result from another immunological assay, such as the MIADA assay. We have developed the high throughput MIADA assay to identify different subtypes of novel influenza A infection^32–34^. In combination with microneutralization assays, MIADA offers a high throughput option for the initial screening during large scale serosurveys. However, it should be emphasized that due to the complex immunity in the population and cross-reactivity from past exposure to seasonal influenza, binding antibody results must be verified with microneutralization assays and HI assays using virus antigenically closely related to the viruses that cause the outbreak, and additional serum adsorption may be needed to confirm true seropositivity to A(H5) ^33,35,36^.

Our study has limitations, first, the clinical protective functions of the cross-reactive NA and HA stalk antibodies to A(H5N1) viruses in humans need to be further understood; second, although we assessed multiple antibody targets; other adaptive immunity, such as cell mediated immunity that can also be associated with reduction of influenza disease severity^13^, were not assessed here.

Host immunity is a critical parameter in assessing population susceptibility to influenza virus infections.

The Centers for Disease Control (CDC) uses the Influenza Risk Assessment Tool (IRAT) ^37^ to evaluate the pandemic potential for a novel influenza A virus. Population immunity is one of the risk elements used in the IRAT. Our study provides additional data for A(H5N1) pandemic risk assessment. Further studies are needed to better understand the immune responses to A(H5N1) viruses in humans and to what extent pre-existing immunity can deter infection and/or lessen disease severity. Continued surveillance is essential to closely monitor A(H5) viruses for pandemic preparedness.

## Materials and Methods

### Human sera

A total of 1794 sera collected from 723 participants in the US were used in the study. Details of the sera collection are described in Table 1.

In an ongoing longitudinal population immunity study, 489 participants from all 10 US HHS regions and 8 age ranges (0-10 yrs, 11-20 yrs, 21-20 yrs, 31-40 yrs, 41-50 yrs, 51-60 yrs, 61-70 yrs, and > 70 yrs) were enrolled in 3 waves from November 2021 to February 2023 (prior to the current A(H5) outbreak in the US) and provided 1467 sera (Table 1 and Fig 1a-b) for analysis.

Seasonal influenza vaccination, infections, and control sera from the 2023-24 season since the outbreak of 2.3.4.4b A(H5) virus in dairy cattle and poultry were also evaluated. For influenza vaccination, pre- (S1) and post- (S2) vaccination sera collected through a CDC contract from pediatric (0.5-2.8 yrs, n=19), adult (20-48 yrs, n=20), and older adult (65-92 yrs, n=20) participants who received quadrivalent inactivated influenza vaccines (IIV4) in 2023-24 were analyzed. For seasonal influenza A infection, sera were collected in the US Flu VE network ^38^ from persons exhibiting acute respiratory illness (ARI) during October 2023 to February 2024 from reverse transcription polymerase chain reaction (RT-PCR) confirmed influenza A(H1N1)pdm09 infection (21 pairs and 41 S1 only), A(H3N2) virus infection (13 pairs and 22 S1 only), and negative cases (n=78) (Table 1).

Acute sera were collected from all participants within 7 days post-symptom onset; for those with RT-PCR confirmed A(H1N1)pdm09 or A(H3N2) virus infections, convalescent sera were collected 21-28 days post-symptom onset.

The use of sera was approved by the Centers for Disease Control and Prevention (CDC) Human Subjects Research Determination. The study was reviewed by CDC and conducted consistent with applicable federal law and CDC policy ^39–43^.

### Multiplex influenza antibody detection assay (MIADA)

In this study, 12-plex and 28-plex cocktails were used that included influenza HA ectodomain (Ec), HA globular head (GH) from A(H1N1), A(H3N2), A(H5N1), influenza B viruses, group 1 and group 2 HA stalks; N1 and N2 NAs, influenza A nucleoprotein (NP), and a protein A/G control, details of the antigens used are described in Extended Table 1 ^44,45^. Antigens were coupled with Bio-Plex Pro™ Magnetic COOH beads. Linearity of the antibody levels was first assessed by serial 2-fold diluted curves of representative sera to determine optimal dilution. Then the 1:2000-diluted human sera were used in the assay followed by incubation with the MIADA antigen cocktail on Mini orbital shaker at room temperature. The secondary antibody of phycoerythrin-conjugated goat F(ab’)2 anti-human pan Immunoglobulin (PE-Pan Ig) reporters were used, and results were analyzed by a Luminex MAGPIX reader to quantify MFIs ^46^.

### Influenza viruses, molecular and structural analyses

Wild type HPAI 2.3.4.4b A(H5N1) virus isolated from the first dairy cattle outbreak human case A/Texas/37/2024^6^ was used in the analyses, additional 2.3.4.4b A(H5N1) virus: A/American wigeon/South Carolina/22-000345-001/2021 (AW/2021) wild type and candidate vaccine virus (CVV), and wild type A/gyrfalcon/Washington/410886/2014 H5N8 (GF/2014) were also used in the analysis. A(H5N1) viruses were propagated in 10–11 day old embryonated eggs. 2023-2024 seasonal A(H1N1)pdm09 virus: A/Wisconsin/67/2022 and A(H3N2) virus: A/Massachusetts/18/2022 were also used. Seasonal A(H3N2) and A(H1N1)pdm09 viruses were propagated in Madin-Darby Canine Kidney (MDCK)-SIAT1 cells.

All studies using HPAI A(H5Nx) viruses were conducted in Biosafety Level 3 Enhanced (BSL3E) laboratories, studies using seasonal viruses were conducted in Biosafety Level 2 (BSL2) laboratories.

Virus stocks used in the study were sequenced. HA head, stalk and NA sequences were analyzed using the BioEdit software (https://bioedit.software.informer.com). HA and NA structural models were generated using PyMOL (Molecular Graphics System).

### Hemagglutination Inhibition (HI) assay using horse erythrocytes

HI assays to detect antibody responses to A(H5) viruses were performed with a modified HI assay using horse erythrocytes as previously described^47,48^. In brief, sera were heat inactivated for 30 min at 56 °C and then tested for non-specific agglutinins and adsorbed with horse erythrocytes as needed. Sera were then treated with receptor-destroying enzyme (RDE) for 18–20 h at 37 °C to remove any non-specific inhibitors that may have been introduced during hemadsorption, followed by 56 °C heat inactivation of RDE for 30 min prior to the HI assay.

Sera were serially diluted two-fold and incubated for 30 min with 4 hemagglutination units per 25 μL of virus, and then incubated with 1% horse erythrocytes for 60 min. HI titer was defined as the reciprocal of the last dilution of serum that completely inhibited hemagglutination. Antibody titer <10 (initial dilution) was reported as 5 for statistical analysis.

### Microneutralization (MN) Assays

MN assays were performed as previously described^47–49^. Heat inactivated human sera were serially diluted 2-fold and incubated with one hundred 50% tissue culture infection dose (TCID50) of influenza viruses. The virus-sera mixture was used to infect 1.5 × 10^4^/well MDCK cells and incubated overnight. The plates were fixed with cold 80% acetone and the presence of viral nucleoprotein was quantified by enzyme linked immunosorbent assay (ELISA). Microneutralization titers were defined as the reciprocal of the highest serum dilution that showed 50% neutralization. Antibody titer <10 (initial dilution) was reported as 5 for statistical analysis.

### Generation of H6Nx reverse genetic viruses with mismatched H6 HA and target NA

Reverse genetics (RG) influenza viruses were generated with a mismatched H6 HA (from A/turkey/Massachusetts/3740/1965, GISAID: EPI407901) and target NA: N1 from 2.3.4.4b A (H5N1) virus A/Texas/37/2024 (GISAID: EPI3171486), N1 from seasonal A(H1N1)pdm09 A/Wisconsin/588/2019 (GISAID: EPI2413538) and N2 from seasonal A(H3N2) A/Darwin/6/2021 (EPI3009099). Briefly, HA and NA genes of interest and 6 internal genes from A/Puerto Rico/8/1934 (H1N1) were synthesized and cloned into the transcription plasmid pCIPolISapIT. Plasmids containing the 8 gene segments were transfected into 293T cells at 1 µg each using Lipofectamine 2000 to produce recombinant virus. Recombinant virus was propagated in 9-11 day-old embryonated hen’s eggs and the genome was confirmed by sequencing. Virus stocks were stored at −80 °C until analysis.

The RG viruses do not have gain of function. Studies using H6Nx RG viruses were conducted in Biosafety II Enhanced Laboratories.

### Enzyme-linked Lectin Assay (ELLA)

Functional NA inhibition antibodies (NAI) were detected using H6Nx RG viruses by the ELLA assay^50^. Sera were first heat inactivated. Serial two-fold diluted sera were then incubated with H6Nx RG viruses in 96-well plates coated with fetuin for 16–18 h. Horse radish peroxidase (HRP)-labeled peanut agglutinin (lectin) was added to the wells and incubated for 2h, followed by washing and the addition of substrate to reveal enzymatic cleavage of fetuin by viral NA. NAI titers were calculated as the reciprocal of the highest dilution with at least 50% inhibition of NA activity. ELLA assays were conducted in Biosafety II Enhanced Laboratories.

### Statistical analysis

Antibody responses comparing pre- vs post-vaccination and acute vs convalescent sera were analyzed using two-tailed *t* tests, *p*<0.05 is considered statistically significant. Statistical analyses were performed using GraphPad Prism 8.

### Data availability

Data generated in this study are provided in the manuscript. Additional raw data from the study are available from the corresponding author Dr. Min Z. Levine upon reasonable request.

## Author contribution

M.Z. L conceived the study; M.Z, L. and Z.L. designed the study; Z.L., F.L., Y.J., S.J., C.H., F.L.G., W.T., P. C., I.Y., C.T.D., J.S., T.T. acquired the data. N.S., A.K., provided sera. M.Z.L. analyzed the data and interpreted the results. M.Z.L., Z.L., F.L. wrote the manuscript. M.Z.L supervised the study. All co-authors were involved in the manuscript preparation process for important intellectual content.

## Acknowledgements

We thank the investigators from US Flu VE network sites for providing the acute and convalescent sera collected from seasonal influenza infections and influenza-negative controls during 2023-24. We thank Ms. Shelly Zimbric, staff from the University of Wisconsin and the study sites of the influenza population immunity study for collecting the 2021-23 population immunity sera. We also thank Xiaonan Sun, Makeda Kay, Sneha Joshi and Hao Zhang from the influenza division, CDC for sample and data management.

## Funding and disclaimer

This study is funded by Centers for Disease Control and Prevention (CDC). The collection of population immunity sera was funded under contract: 200-2016-91800 Safety and Healthcare Epidemiology Prevention Research Development (SHEPheRD).

Opinions, findings, and conclusions reported by the authors are strictly their own and are in no way meant to represent the opinion, views, policies or the official position of CDC and U.S. Department of Health and Human Services.

## Additional Information

Correspondence and requests for materials should be addressed to Dr Min Z. Levine

## Extended Data Figures and Tables

**Extended Fig. 1:**
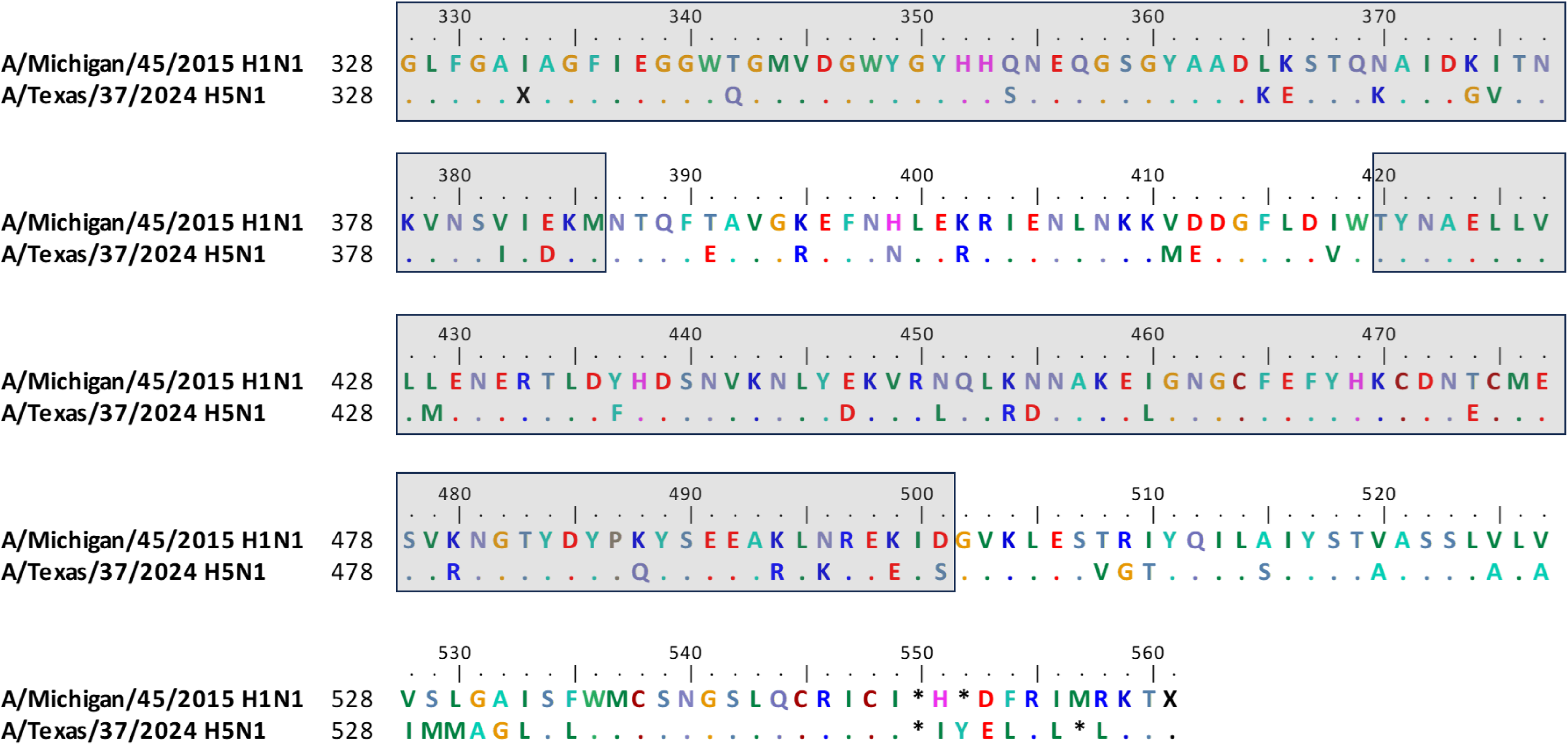
Amino acid differences between the H1N1 HA stalk and H5N1 HA stalk antigens. HA stalk sequences from the A(H1N1) and A(H5N1) viruses were analyzed using ClustalW multiple alignment in BioEdit software. HA stalk residuals 328-386 and 420-501 (shown in the grey boxes) were expressed in the HA stalk construct using methods previously described^14^. Identical amino acids referred to the HA stalk sequence of A/Wisconsin/67/2022 H1N1 virus were presented as dots. Amino acid homology of the expressed HA stalk residuals between the two viruses is 84%.

**Extended Fig. 2:**
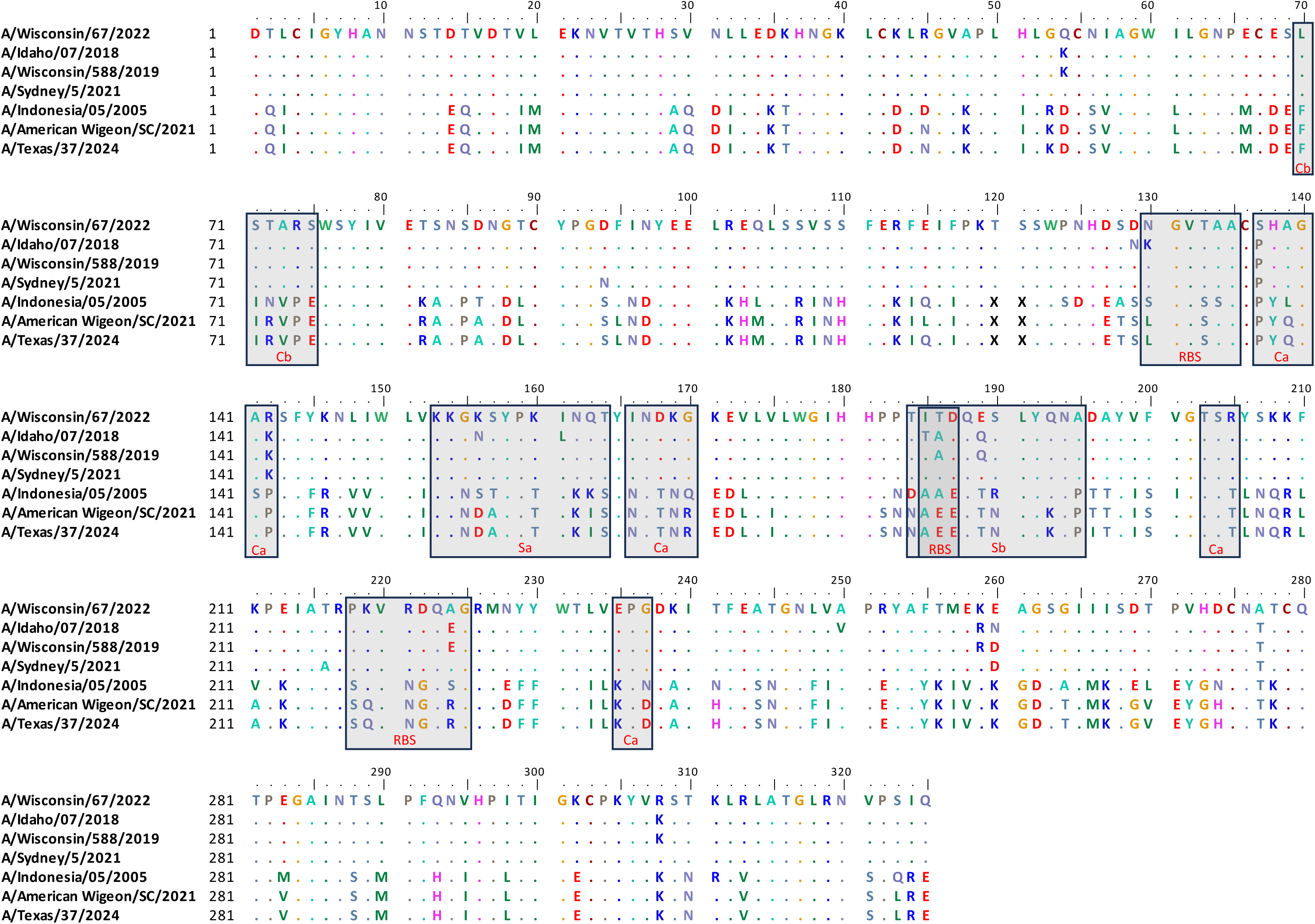
Amino acid differences between the HA head of A(H5N1) and A(H1N1)pdm09 viruses analyzed. HA1 sequences from the H1N1 and H5N1 viruses were analyzed using ClustalW multiple alignment in BioEdit software. Conventional antigenic sites (Ca, Cb, Sa, Sb) and receptor binding sites (RBS) on the H1 HA head domain were highlighted in the grey boxes. Identical amino acids referred to the HA1 sequence of A/Wisconsin/67/2022 H1N1pdm09 virus were presented as dots.

**Extended Fig. 3:**
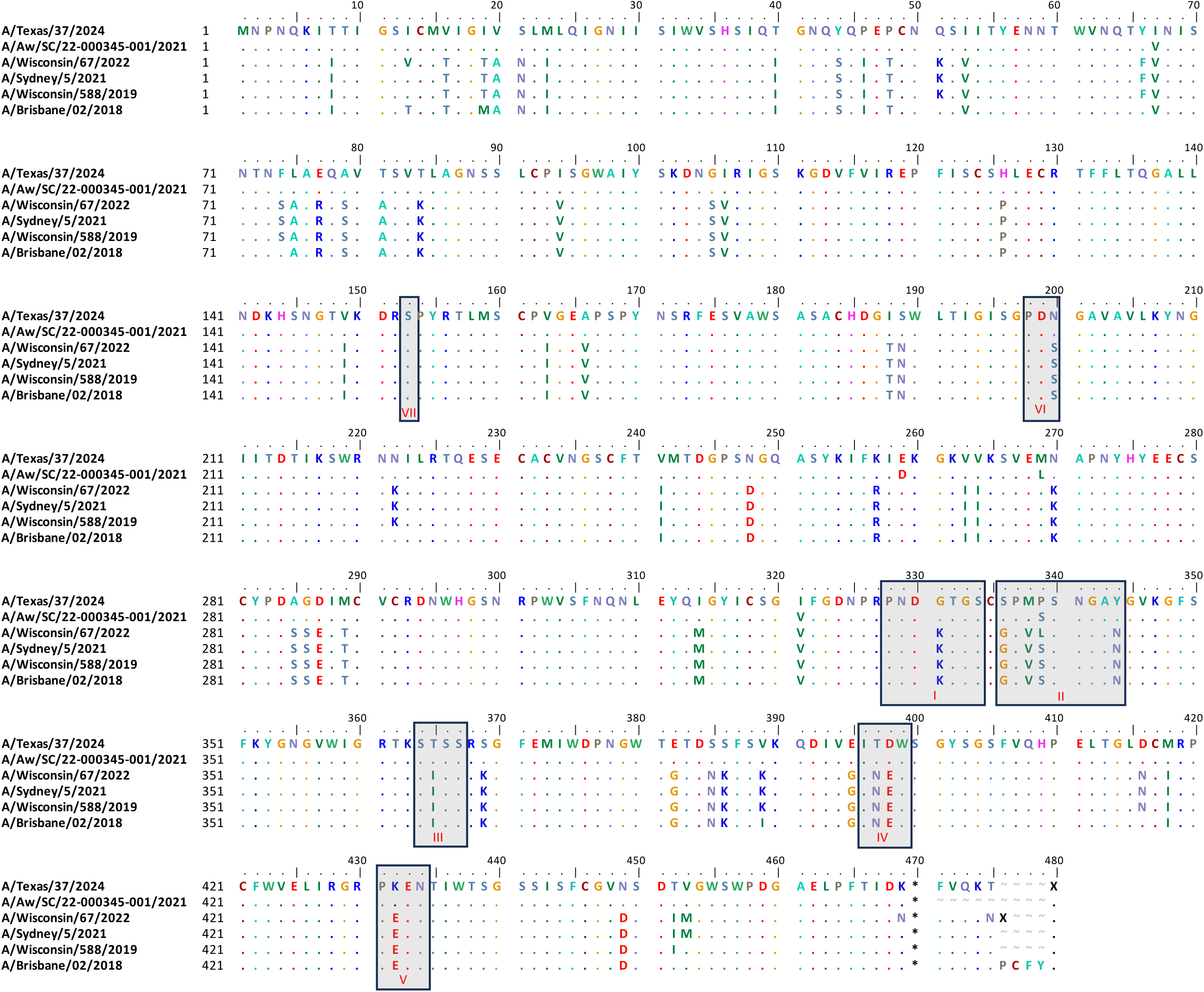
Amino acid differences among the N1 NA of the A(H5N1) and A(H1N1)pdm09 viruses analyzed. NA sequences from the H1N1 and H5N1 viruses were analyzed using ClustalW multiple alignment in BioEdit software. Putative NA antigenic sites (I to VII) were highlighted in the grey boxes. Identical amino acids referred to the NA

**Extended Fig. 4:**
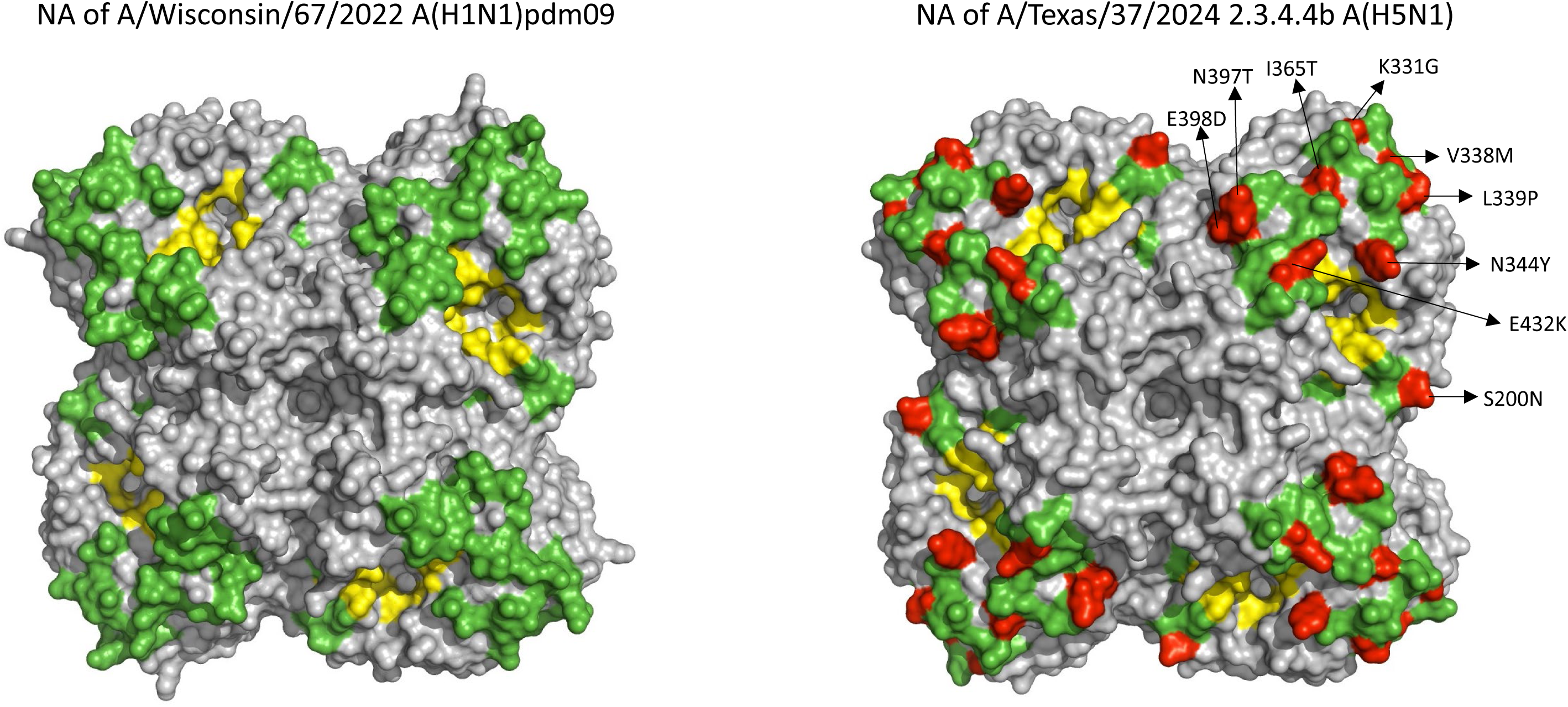
Comparison of amino acid residues in the potential NA antigenic sites between A/Wisconsin/67/2022 H1N1 pdm09 and A/Texas/37/2024 H5N1 viruses. The NA tetramers were modeled with PyMOL version 2.1.1 from A/Michigan/45/2015 H1N1 pdm09 (PDB: 7S0I) for A/Wisconsin/67/2022 H1N1 pdm09 NA and from A/American green-winged teal/Washington/195750/2014 H5N1 (PDB: 5HUG) for A/Texas/37/2024 H5N1 NA respectively. Putative NA antigenic sites were shown in green, of which 10 amino acid differences (at site 200, 331, 336, 338, 339, 344, 365, 397, 398, 432) between the two NA sequences were highlighted in red on the modeling of A/Texas/37/2024 H5N1 virus. G336S change was underneath the 338 and 339 sites and was not visible from the top view of this tetramer. The amino acid residues in the enzyme active site were presented in yellow.

**Extended Table S1.** Antigens used in the multiplex influenza antibody detection (MIADA) assay.

| Assays<br>(N-plex) | Antigen group | Antigen | Virus strain | Type (subtype or lineage) | HA<br>group | Egg or Cell-<br>origin | Ecto/GH<br>HA1 | Source | GISAIID/GenBank<br>Accession No. |
| --- | --- | --- | --- | --- | --- | --- | --- | --- | --- |
| 12-plex | H1 HA head | H1. WI.19 GH | A/Wisconsin/588/2019 | A(H1N1)pdm09 | 1 | Cell | GH HA1 | CDC | EPI1661758 |
| 12-plex |  | H1.Syd.21 GH | A/Sydney/5/2021 | A(H1N1)pdm09 | 1 | Cell | GH HA1 | CDC | EPI1957293 |
| 12-plex | H3 HA head | H3.Cam.20 GH | A/Cambodia/e0826360/2020 | A(H3N2) | 2 | Cell | GH HA1 | CDC | EPI1837753 |
| 12-plex |  | H3.Dar.21 GH | A/Darwin/6/2021 | A(H3N2) | 2 | Cell | GH HA1 | CDC | EPI1857216 |
| 12-plex | H5 HA head | H5.AW.SC.21 GH | A/American wigeon/South Carolina/22-000345-001/2021 | A(H5N1) | 2 | Cell | GH HA1 | CDC | EPI1985910 |
| 12-plex | Group 1 HA stalk | H1. MI.15 Stalk | A/Michigan/45/2015 | A(H1N1)pdm09 | 2 | Cell | HA Stalk | CDC | EPI662594 |
| 12-plex | Group 2 HA stalk | H3.Sin.16 Stalk | A/Singapore/INFIMH-16–0019/2016 | A(H3N2) | 2 | Cell | HA Stalk | CDC | EPI780183 |
| 12-plex | B HA head | B-V-WA.19 GH | B/Washington/02/2019 | B/Victoria lineage | N/A | Clinical | GH HA1 | CDC | EPI1368874 |
| 12-plex |  | B-V-Aus.21 GH | B/Austria/1359417/2021 | B/Victoria lineage | N/A | Cell | GH HA1 | CDC | EPI1845793 |
| 12-plex |  | B-Y-Phu.13 GH | B/Phuket/3073/2013 | B/Yamagata lineage | N/A | Cell | GH HA1 | CDC | EPI529345 |
| 12-plex | NP | A-NP | A/Brisbane/10/2007 | A(H3N2) | N/A | Egg | NP | IRR | EPI176962 |
| 12-plex | Protein A/G | Protein A/G | N/A | N/A | N/A | N/A | N/A | ThermoFisher | N/A |
| 28-plex | H1 HA head | H1.ID.18 GH | A/Idaho/07/2018 | A(H1N1)pdm09 | 1 | Cell | GH HA1 | CDC | EPI1206974 |
| 28-plex |  | H1. WI.19 GH | A/Wisconsin/588/2019 | A(H1N1)pdm09 | 1 | Cell | GH HA1 | CDC | EPI1661758 |
| 28-plex |  | H1.Syd.21 GH | A/Sydney/5/2021 | A(H1N1)pdm09 | 1 | Cell | GH HA1 | CDC | EPI1957293 |
| 28-plex |  | H1.WI.22 GH | A/Wisconsin/67/2022 | A(H1N1)pdm09 | 1 | Cell | GH HA1 | CDC | EPI2224788 |
| 28-plex | H3 HA head | H3.Cam.20 GH | A/Cambodia/e0826360/2020 | A(H3N2) | 2 | Cell | GH HA1 | CDC | EPI1837753 |
| 28-plex |  | H3.Dar.21 GH | A/Darwin/6/2021 | A(H3N2) | 2 | Cell | GH HA1 | CDC | EPI1857216 |
| 28-plex |  | H3.MA.22 GH | A/Massachusetts/18/2022 | A(H3N2) | 2 | Cell | GH HA1 | CDC | EPI2096148 |
| 28-plex | H5 HA ectodomain | H5.Ind.05 Ec | A/Indonesia/05/2005 HA (H5N1) | A(H5N1) | 1 | Egg | Ecto | CDC | EPI376537 |
| 28-plex |  | H5.Tx.24 Ec | A/Texas/37/2024 (H5N1) | A(H5N1) | 1 | Egg | Ecto | CDC | EPI3171488 |
| 28-plex | H5 HA head | H5.Ind.05 GH | A/Indonesia/05/2005 HA (H5N1) | A(H5N1) | 1 | Egg | GH HA1 | CDC | EPI376537 |
| 28-plex |  | H5.AW.SC.21 GH | A/American wigeon/South Carolina/22-000345-001/2021 | A(H5N1) | 1 | Egg | GH HA1 | CDC | EPI1985910 |
| 28-plex |  | H5.Tx.24 GH | A/Texas/37/2024 (H5N1) | A(H5N1) | 1 | Egg | GH HA1 | CDC | EPI3171488 |
| 28-plex | B HA head | B-V-WA.19 GH | B/Washington/02/2019 | B/Victoria lineage | N/A | Clinical | GH HA1 | CDC | EPI1368874 |
| 28-plex |  | B-V-Aus.21 GH | B/Austria/1359417/2021 | B/Victoria lineage | N/A | Cell | GH HA1 | CDC | EPI1845793 |
| 28-plex |  | B-Y-Phu.13 GH | B/Phuket/3073/2013 | B/Yamagata lineage | N/A | Cell | GH HA1 | CDC | EPI529345 |
| 28-plex | Seasonal N1 | N1 NA.BR 18 | A/Brisbane/02/2018 | A(H1N1)pdm09 | N/A | Cell | N1 NA | Sino Biological US Inc | EPI1799927 |
| 28-plex |  | N1 NA.Wis.19 | A/Wisconsin/588/2019 | A(H1N1)pdm09 | N/A | Cell | N1 NA | Sino Biological US Inc | EPI1661757 |
| 28-plex |  | N1 NA.Syd 21 | A/Sydney/5/2021 | A(H1N1)pdm09 | N/A | Cell | N1 NA | Sino Biological US Inc | EPI2413554 |
| 28-plex |  | N1 NA.Wis.22 | A/Wisconsin/67/2022 | A(H1N1)pdm09 | N/A | Cell | N1 NA | Sino Biological US Inc | EPI2224787 |
| 28-plex | Seasonal N2 | N2 NA.Cam 20 | A/Cambodia/e0826360/2020 | A(H3N2) | N/A | Cell | N2 NA | Sino Biological US Inc | EPI1837752 |
| 28-plex |  | N2 NA.Dar.21 | A/Darwin/6/2021 | A(H3N2) | N/A | Cell | N2 NA | Sino Biological US Inc | EPI1925254 |
| 28-plex |  | N2 NA.MA.22 | A/Massachusetts/18/2022 | A(H3N2) | N/A | Cell | N2 NA | CDC | EPI2096147 |
| 28-plex | N1 (H5N1) | N1.H5.AW.SC.21 | A/American wigeon/South Carolina/22-000345-001/2021 | A(H5N1) | N/A | Cell | N1 NA | CDC | EPI1985912 |
| 28-plex |  | N1 NA H5.Tx.24 | A/Texas/37/2024 (H5N1) | A(H5N1) | N/A | Egg | N1 NA | CDC | EPI3171486 |
| 28-plex | Group 1 HA stalk | H1. MI.15 Stalk | A/Michigan/45/2015 | A(H1N1)pdm09 | 1 | Cell | HA Stalk | CDC | EPI662594 |
| 28-plex | Group 2 HA stalk | H3.Sin.16 Stalk | A/Singapore/INFIMH-16–0019/2016 | A(H3N2) | 2 | Cell | HA Stalk | CDC | EPI780183 |
| 28-plex | NP | A-NP | A/Brisbane/10/2007 | A(H3N2) | N/A | Egg | NP | IRR | EPI176962 |
| 28-plex | Protein A/G | Protein A/G | N/A | N/A | N/A | N/A | N/A | ThermoFisher | N/A |

GH: Globular head

**Extended Table S2:** Microneutralization assays and hemagglutination inhibition assays against wild type 2.3.4.4b HPAI H5N1 viruses to confirm seropositivity of 2021-23 sera with high MFIs to 2.3.4.4b H5N1 virus HA.

| Sera | A/American wigeon/SC/22-00345-001/2021 H5N1 HA head | Wild type A/American wigeon/SC/22-00345-001/2021 H5N1 virus |  | Wild type A/Texas/37/2024 H5N1 virus |  |
| --- | --- | --- | --- | --- | --- |
|  | MFI in MIADA (Binding antibody ) | Neutralizing antibody* | HI antibody* | Neutralizing Antibody* | HI Antibody* |
| 1 | 3889 | 5 | 5 | 5 | 5 |
| 2 | 1239 | 5 | 5 | 5 | 7 |
| 3 | 1159 | 5 | 5 | 5 | 5 |
| 4 | 1139 | 5 | 5 | 5 | 7 |
| 5 | 1072 | 5 | 5 | 5 | 7 |
| 6 | 1025 | 5 | 5 | 5 | 5 |
| 7 | 852 | 5 | 5 | 5 | 7 |
| 8 | 842 | 5 | 5 | 5 | 7 |
| 9 | 807 | 5 | 5 | 5 | 7 |
| 10 | 785 | 5 | 5 | 5 | 5 |
| 11 | 716 | 5 | 5 | 5 | 5 |
| 12 | 678 | 5 | 5 | 5 | 7 |

\*: Sera were tested at pre-dilution 1:10 in MN and HI assays, titers less than 10 are reported as 5. Reported titers are geometric mean titers of 2 replicates

**Extended Table S3:** Percent amino acid homology of HA1 sequences of A(H5N1) and recent seasonal A(H1N1)pdm09 viruses.

|  | A/Texas/37/2024 H5N1<br>(2.3.4.4b) | A/American wigeon/2021 H5N1<br>(2.3.4.4b) | A/Indonesia/05/2005<br>H5N1 (2.1) | A/Wisconsin/67/2022<br>H1N1 | A/Sydney/5/2021<br>H1N1 | A/Wisconsin/588/2019<br>H1N1 | A/Idaho/07/2018<br>H1N1 |
| --- | --- | --- | --- | --- | --- | --- | --- |
| A/Texas/37/2024 H5N1 (2.3.4.4b) | N.A | 99 | 88 | 53 | 53 | 53 | 53 |
| A/American wigeon/2021 H5N1 (2.3.4.4b) | 99 | N.A | 88 | 53 | 53 | 53 | 53 |
| A/Indonesia/05/2005 H5N1 (2.1) | 88 | 88 | N.A | 52 | 53 | 53 | 53 |
| A/Wisconsin/67/2022 H1N1 | 53 | 53 | 52 | N.A | 98 | 97 | 95 |
| A/Sydney/5/2021 H1N1 | 53 | 53 | 53 | 98 | N.A | 98 | 95 |
| A/Wisconsin/588/2019 H1N1 | 53 | 53 | 53 | 97 | 98 | N.A | 98 |
| A/Idaho/07/2018 H1N1 | 53 | 53 | 53 | 95 | 95 | 98 | N.A |

N/A: non applicable

**Extended Table S4:**
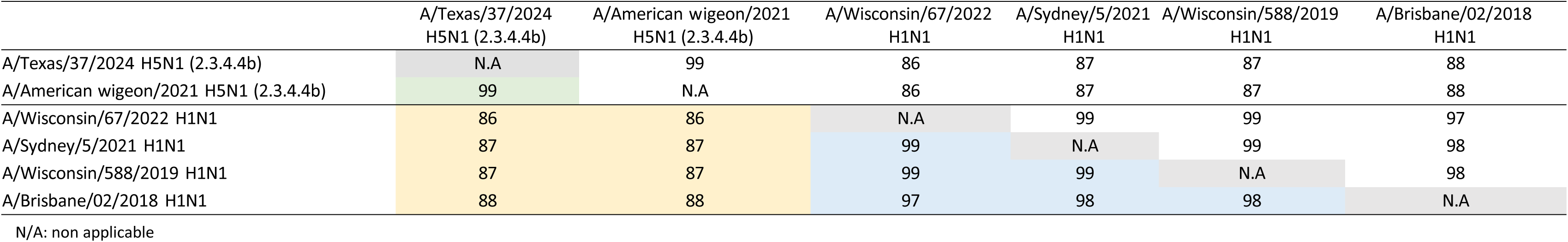
Percent amino acid homology of NA sequences of A(H5N1) and recent seasonal A(H1N1)pdm09 viruses.

|  | A/Texas/37/2024<br>H5N1 (2.3.4.4b) | A/American wigeon/2021<br>H5N1 (2.3.4.4b) | A/Wisconsin/67/2022<br>H1N1 | A/Sydney/5/2021<br>H1N1 | A/Wisconsin/588/2019<br>H1N1 | A/Brisbane/02/2018<br>H1N1 |
| --- | --- | --- | --- | --- | --- | --- |
| A/Texas/37/2024 H5N1 (2.3.4.4b) | N.A | 99 | 86 | 87 | 87 | 88 |
| A/American wigeon/2021 H5N1 (2.3.4.4b) | 99 | N.A | 86 | 87 | 87 | 88 |
| A/Wisconsin/67/2022 H1N1 | 86 | 86 | N.A | 99 | 99 | 97 |
| A/Sydney/5/2021 H1N1 | 87 | 87 | 99 | N.A | 99 | 98 |
| A/Wisconsin/588/2019 H1N1 | 87 | 87 | 99 | 99 | N.A | 98 |
| A/Brisbane/02/2018 H1N1 | 88 | 88 | 97 | 98 | 98 | N.A |

N/A: non applicable

